# Impact of LLM Assistance on Physician Decision-Making: A Multi-Country Randomized Controlled Trial^∗^

**DOI:** 10.1101/2025.08.08.25333272

**Authors:** Nicholas Rounding, Luthfi Saiful Arif, Janine Berg, Jochen Cals, Diederik De Boer, Eefje De Bont, Sander Dijksman, Ardi Findyartini, Didier Fouarge, Marie-Christine Fregin, Pawel Gmyrek, Nadia Greviana, Ralph Leijenaar, Soraiya Manji, Annastacia Mbithi, Norah Obungu, Arierta Pujitresnani, Roselyter Rianga, Diantha Soemantri, Sairabanu Mohamed Rashid Sokwalla, Sanne Steens, Lucia Velasco, Ardy Wildan, Prasandhya Astagiri Yusuf, Mark Levels

## Abstract

Disparities in the quality of healthcare persist globally, with poor-quality care contributing significantly to preventable mortality, particularly in low- and middle-income countries. While digital technologies, including generative artificial intelligence (AI), hold promise for improving clinical decision-making, their global effectiveness and potential to mitigate cross-country variation remain underexplored. We conducted a parallel-group randomized controlled trial across three economically diverse countries—Indonesia, Kenya, and the Netherlands—to evaluate the impact of large language model (LLM) access on physician performance using standardized clinical vignettes. Physicians (N=249) were randomly assigned to either a control group or an intervention group with access to GPT-4o. Results showed that LLM access significantly improved clinical performance, with the largest effect in Kenya (18%, 95% CI: 12.7 to 23.2, p*<*0.001), followed by Indonesia (10.7%, 95% CI: 5.7 to 15.7, p*<*0.001) and the Netherlands (7.2%, 95% CI: 3.7 to 10.7, p*<*0.001). Notably, LLM access reduced cross-country performance disparities, particularly between Kenya and the Netherlands. However, distributional effects varied, with increased score dispersion in Indonesia and reduced variation in Kenya. Higher LLM usage was associated with greater performance gains, though some physicians without access outperformed those with access, suggesting that effective use depends on individual engagement. Our findings demonstrate that LLMs can enhance clinical performance across diverse settings while potentially narrowing global inequalities in care quality. Further research should explore mechanisms of effective LLM integration and long-term impacts on real-world clinical practice.

## 1 Introduction

Over the past decades many regions have expanded basic health coverage, yet disparities in the quality of care remain a pervasive global challenge [15]. Poor-quality of care now contributes more to mortality than lack of access, with particularly severe consequences in low- and middle-income countries where 60% of deaths from treatable and preventable conditions arising from poor-quality care with the remainder arising from non-utilization of healthcare [33]. Inequalities in the quality of care have been recorded within and across countries [6, 16, 44]. Improving the quality of physician care could eliminate millions of unnecessary deaths every year and reduce inequities in care provision between the Global North and South, and between population groups within a country. The value of digital technologies to improve the provision of healthcare, and the quality of physician care, is increasingly being acknowledged by healthcare providers and governments [3, 17, 54, 56].

Innovations in Generative AI, i.e. Large Language Models (LLMs), have been proposed as tools that can augment the provision of healthcare by aiding physicians in their work [5, 21, 34, 52]. LLMs have shown potential in key clinical tasks, including clinical reasoning and generating differential diagnoses, and have demonstrated strong performance in simulated clinical environments [11, 14, 37, 40, 50, 53]. Recent studies suggest that LLMs could help improve the quality of care by augmenting physician clinical decision-making, including both diagnosis and management tasks [18, 24, 25, 32]. Whether physicians will be directly replaced by AI or not is debated, research suggests that the exposure of physicians to task replacement by LLMs is relatively low suggesting augmentation is more likely in the immediate future [19, 22]. Given this, it is imperative to confirm and generalize the results of similar trials in global contexts, where results could differ due to cultural differences in clinical reasoning [20, 31], technology acceptance [39] and different regulatory and organizational contexts. Furthermore, the effect of LLMs on cross-country variation in physician performance has been unexplored.

To investigate whether LLM use is globally effective and the extent to which it can mitigate inequalities in physician performance, we designed a parallel group randomized controlled trial to evaluate the effectiveness of an LLM (GPT-4o) in improving physician clinical performance on vignettes. We administered clinical vignettes[47] in simulated primary care scenarios across three economically and geographically diverse countries: Indonesia, Kenya, and the Netherlands. These countries represent distinct income strata, upper-middle-income, lower-middle-income, and high-income, respectively, offering a broader perspective on regional and economic differences in healthcare delivery. Clinical vignettes have been used to assess the augmentative abilities of LLMs in other studies [18, 24, 25]. Performance on clinical vignettes is a validated measure of the quality of physician clinical practice and care, comparable to the gold standard of standardized patients [47]. Physicians’ answers in our vignettes are graded against detailed rubrics built from context specific, evidence-based, best practice guidelines. Such guidelines have been demonstrated to improve quality of care globally [26, 27, 36, 58]. By comparing the outcomes of physicians with and without access to the LLM in three countries, we analyze how LLM access affects variation in physician performance within and across countries.

## 2 Results

We recruited 249 resident physicians: 81 in Indonesia, 60 in Kenya, and 108 in the Netherlands. Data were collected in August–September 2024 (Netherlands), November 2024 (Indonesia), and January 2025 (Kenya). Table 1 reports baseline characteristics. In Indonesia, all participants were internal medicine residents. In Kenya, 45% were internal medicine residents, while the remaining 55% were either first-year residents in other specialties (i.e., surgery, anesthesiology, and pediatrics) or post-internship pre-residency medical officers referred to as Senior House Officers in Kenya. We refer to this group collectively as the non-internal medicine subgroup throughout. In the Netherlands, all participants were family medicine specialists, 83% were residents and 17% were attending physicians. The mean number of years since beginning medical education was 13 in Indonesia, 11.8 in Kenya, and 13.9 in the Netherlands Participants were randomly assigned to either a control or intervention group, with the latter receiving access to an LLM (GPT-4o). They were administered the same 4 clinical vignettes in a randomized order, in English, via the Qualtrics survey environment.

**Table 1:**
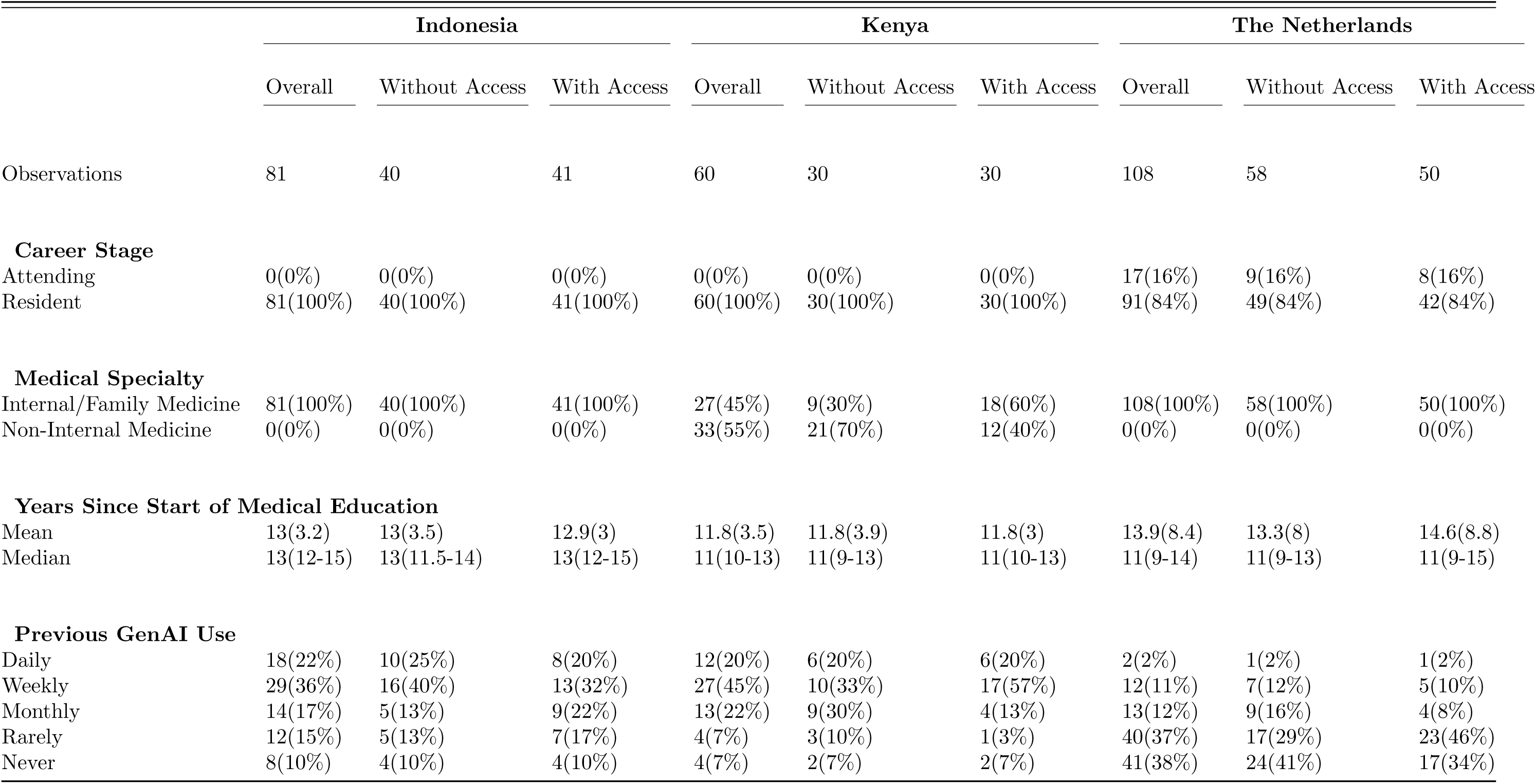
Baseline Characteristics.

### 2.1 Effect of LLM Access on Vignette Performance

Figure 2 displays the distributions of our samples comparing physicians with and without LLM access, showing higher medians, interquartile ranges (IQRs), and boxplot tails in each country. Table 2 presents linear regression model estimates for the effect of LLM access on physician average vignette scores. We observed the largest difference between physicians with and without LLM access in the Kenyan sample 18% (95% CI: 12.7 to 23.2, p<0.001), followed by Indonesia 10.7% (95% CI: 5.7 to 15.7, p<0.001) and the Netherlands 7.2% (95% CI: 3.7 to 10.7, p<0.001). We address the effect of the Kenyan non-internal medicine resident subgroup in Supplement 2, which found a larger effect of 20.5% (95% CI: 13.5 to 27.5, p<0.001) for the non-internal medicine subgroup. We found a correspondingly smaller effect of 10.6% (95% CI: 3.4 to 17.9, p=0.006) for the internal medicine residents, is similar in magnitude to that found in Indonesia and the Netherlands.

**Figure 1:**
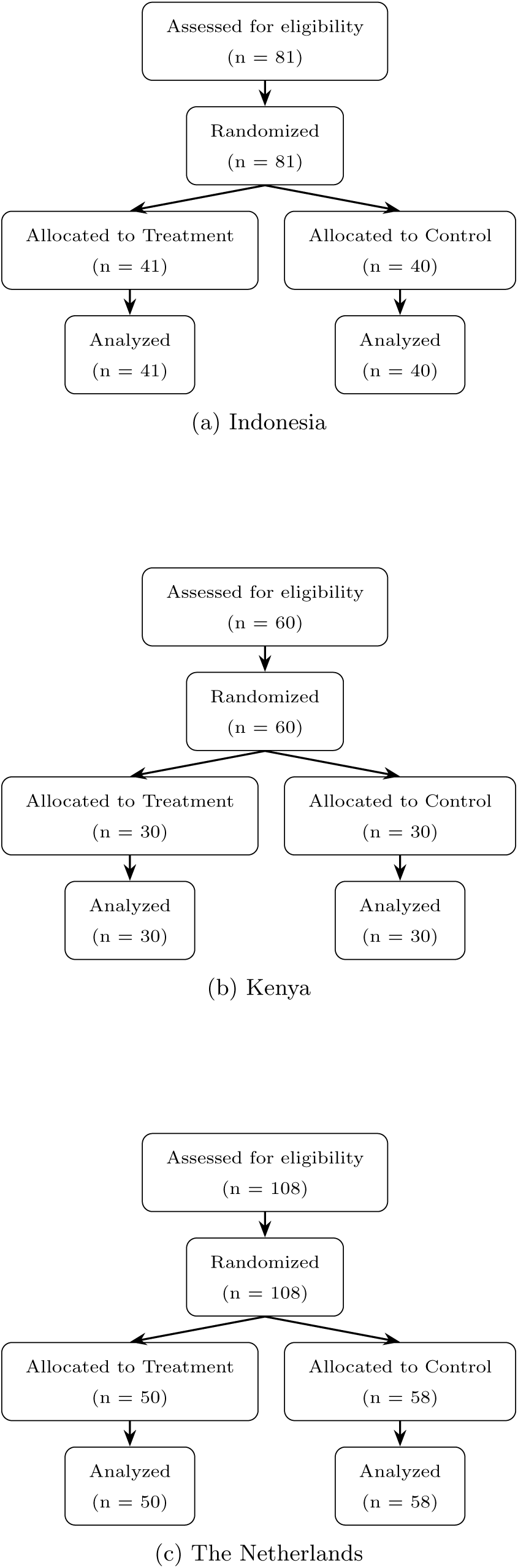
Participant Flow Diagrams by Country

**Figure 2:**
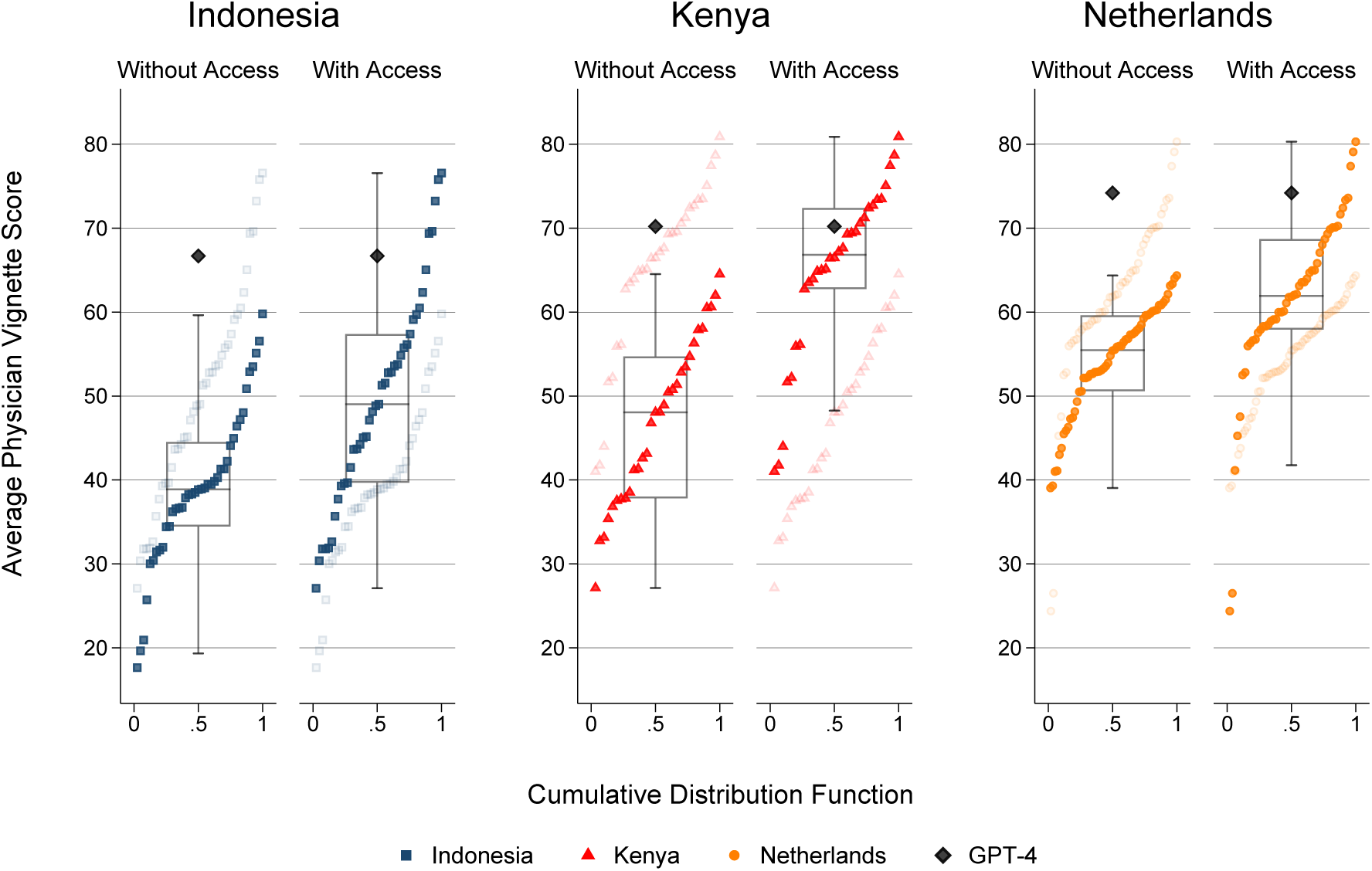
Comparison of Average Physician Vignette Scores Across Countries With and Without LLM Access *Note:* The figure shows the distribution of average physician vignette scores across the three countries, grouped by Without and With LLM access. Boxplots indicate score distributions: the boxes span the interquartile range (IQR), and whiskers extend to the minimum or maximum values within 1.5×IQR. Individual scores are shown as squares, dots or triangles, and overlaid cumulative distribution functions (CDFs) provide a smoothed view of score distributions within each group. The GPT-4o benchmark score is included in both the With and Without access groups as a reference.

**Table 2:** Comparison of Average Physician Vignette Scores Across Countries With and Without LLM access.

|  |  | Without Access | With Access | Mean Differences (CI 95%) |
| --- | --- | --- | --- | --- |
| **Panel A-Within Country Effects** |  |  |  |  |
| Indonesia | Observations | 40 | 41 |  |
|  | Mean(SD) | 39.2(9.5) | 49.9(12.9) | 10.7(5.7-15.7), p<0.001 |
|  | Median(IQR) | 38.9(34.4-44.5) | 49(39.7-57.4) |  |
|  | Min-Max Difference (Min-Max) | 42.2(17.6-59.8) | 49.5(27.1-76.6) |  |
|  | Kolmogorov-Smirnov |  |  | p<0.001 |
| Kenya | Observations | 30 | 30 |  |
|  | Mean(SD) | 47(10) | 65(10.4) | 18(12.7-23.2), p<0.001 |
|  | Median(IQR) | 48.1(37.8-54.7) | 66.8(62.7-72.4) |  |
|  | Min-Max Difference (Min-Max) | 37.4(27.1-64.5) | 39.9(41-80.9) |  |
|  | Kolmogorov-Smirnov |  |  | p<0.001 |
| Netherlands | Observations | 58 | 50 |  |
|  | Mean(SD) | 54.2(6.5) | 61.4(10.9) | 7.2(3.7-10.7), p<0.001 |
|  | Median(IQR) | 55.5(50.6-59.6) | 61.9(57.9-68.7) |  |
|  | Min-Max Difference (Min-Max) | 25.4(39-64.4) | 55.9(24.4-80.3) |  |
|  | Kolmogorov-Smirnov |  |  | p<0.001 |
| **Panel B - Cross-National Effects** |  |  |  |  |
| Indonesia - Kenya | Mean (95% CI) | 7.8(3.1-12.5),p=0.001 | 15.1(9.6-20.6),p<0.001 | 7.3(0.1-14.5),p=0.05 |
|  | Kolmogorov-Smirnov | p=0.010 | p<0.001 |  |
| Indonesia - Netherlands | Mean (95% CI) | 15(11.6-18.4),p<0.001 | 11.5(6.4-16.5),p<0.001 | -3.5(-9.6-2.5),p=0.25 |
|  | Kolmogorov-Smirnov | p<0.001 | p<0.001 |  |
| Kenya - Netherlands | Mean (95% CI) | 7.2(3.2-11.1),p<0.001 | -3.6(-8.5-1.3),p=0.14 | -10.8(-17- -4.6),p<0.001 |
|  | Kolmogorov-Smirnov | p=0.002 | p=0.04 |  |

We found notable differences in score distributions between physicians with access and physicians without access across countries. In Indonesia, we found a larger Inter-quartile Range (IQR) for physicians with access (17.7%) compared to those without (10.1%). In contrast, for the Kenyan samples the IQR was narrower for those with access (9.7%) compared to those the without (16.9%). We demonstrate in Supplement 2 that the distributional effects were similar for both the internal medicine and non-internal medicine subgroups. In the Netherlands we observed a similar IQR range for those with access (10.8%) to those without (9%). Standard deviations were larger for physicians with access in Indonesia (12.9) compared to those without (9.5) and also for those with access in the Netherlands (10.9) compared to without (6.5). Standard deviations were broadly similar in Kenya.

We also observed differences in the tails of the score distributions, reported on in Panel A of Table 2. In the Indonesian sample, the difference between the lowest and the highest scorers was greater for those with access (49.5%) than without (42.2%). This was even more pronounced for the Netherlands, where the difference for those with access (55.9%) was larger than for those without (25.4%). In Kenya, we detected a similar difference for those with access (39.9%) compared to those without (37.4%).

Importantly, we found that the distributions of physicians with and without access overlapped in all three countries, with the best performing physicians without access outperforming a proportion of physicians with access. Intriguingly, in the Netherlands, we found the lowest scores in the group with access (24.4%).

### 2.2 Effect of LLM Access on Cross-Country Differences

Investigating cross-country differences, we found that physicians without LLM access had the lowest mean score in Indonesia (39.2%), followed by those in Kenya (47%) and in the Netherlands (54.2%). For those with access, we found the lowest mean score in Indonesia (49.9%), followed by the Netherlands (61.4%), and Kenya (65%)

Table 2, Panel B reports on the statistical significance of differences in cross-country differences. We found a significant reduction in the cross-country difference in mean physicians scores between the Kenyan and Dutch samples,-10.8% (95% CI: -17 to -4.6, p<0.001). We found a smaller, but still negative, effect for the difference between the Indonesian and Dutch physicians of -3.5% (95% CI: -9.6 to 2.5, p=0.25) and a corresponding larger difference between the Indonesian and Kenyan physicians of 7.3% (95% CI: 0.1 to 14.5, p=0.05).

Cross-country Kolmogorov–Smirnov tests confirm that the statistical distribution of physician scores were significantly different for each pairwise comparison: Indonesia–Kenya (p=0.01), Indonesia–Netherlands (p<0.001), and Kenya–Netherlands (p=0.002). For physicians with access, we found significant differences for the Indonesia–Kenya (p<0.001) and Indonesia–Netherlands (p<0.001) distributions for those with access. In contrast, only weakly significant differences were found for Kenya–Netherlands (p=0.04).

### 2.3 LLM Vignette Performance

To assess the baseline performance of the LLM alone, we input the vignette cases exactly as they were shown to physicians into GPT-4o exactly once, using no fine-tuned prompting (i.e. zero shot learning). As shown in Figure 2, the model outperformed the highest-scoring physicians who did not have access to an LLM in Indonesia(LLM=66.7%, Best Physician=59.8%), Kenya (LLM=70.2%, Best Physician=64.5%), and the Netherlands (LLM=74.2%, Best Physician=64.4%).

Comparing physicians with access to the LLM, we detect that the LLM outperformed most physicians. However, the highest scoring physicians with access in each country outperformed the LLM score. In Indonesia, the LLM scored 66.7%, putting it below the 90th percentile (69.4%). In Kenya, the LLM scored 70.2%, putting it below the 75th percentile (72.4%). While, in the Netherlands the LLM scored 74.2%, putting it above the 90th percentile (72.9%), but below the 95th percentile (77.4%).

### 2.4 Effect of LLM Usage on Vignette Performance

Participants in the intervention group were provided with LLM access, usage was encouraged but not required. Hence we present an intent-to-treat analysis. To further explore the impact of usage intensity, we classified physicians in the intervention group into High Usage and Low Usage groups, using the median rate of LLM use detected in each country as the threshold. We acknowledge that usage was not randomly assigned and inferences are limited by potential self-selection. Table 3 presents the distribution of participants in each category.

**Table 3:** Comparison of Average Physician Vignette Scores Across Countries with High and Low Usage.

|  |  | Low Usage | High Usage | Mean Differences (CI 95%) |
| --- | --- | --- | --- | --- |
| Indonesia | Observations(%) | 22(54%) | 19(46%) |  |
|  | Mean(SD) | 42.4(10) | 58.5(10.4) | 16.1(9.6-22.5), p<0.001 |
|  | Median(IQR) | 40.6(32.6-49) | 56.1(51.6-69.4) |  |
| Kenya | Observations(%) | 15(50%) | 15(50%) |  |
|  | Mean(SD) | 60.8(11.7) | 69.2(7.2) | 8.4(1.1-15.6), p0.03 |
|  | Median(IQR) | 65.1(51.7-70.6) | 69.4(64.9-73.5) |  |
| Netherlands | Observations(%) | 25(50%) | 25(50%) |  |
|  | Mean(SD) | 57.6(10.3) | 65.1(10.3) | 7.5(1.6-13.3), p0.01 |
|  | Median(IQR) | 58.3(56-65) | 64(61.8-70.3) |  |

Figure 3 displays performance outcomes for High and Low Usage across countries. Due to the small sample sizes, estimates may be more sensitive to outliers and should therefore be interpreted with caution. Nevertheless, average scores were consistently higher for High Usage physicians in all three countries. Table 3 reports the estimated additional effects of high use. In Indonesia, we found that the High Usage sub group scored higher than the Low Usage sub group on our vignettes by 16.1% (95% CI: 9.6 to 22.5, p<0.001). In Kenya, the estimated difference was 8.4% (95% CI: 1.1 to 15.6, p=0.03). In the Netherlands, the difference was 7.5% (95% CI: 1.6 to 13.3, p=0.01). These results indicate that higher levels of LLM use are descriptively associated with higher average performance. However, some physicians in the Low Usage and no access groups achieved higher scores than some in the High Usage group, suggesting that access to an LLM alone is not a necessary condition for high-quality performance.

**Figure 3:**
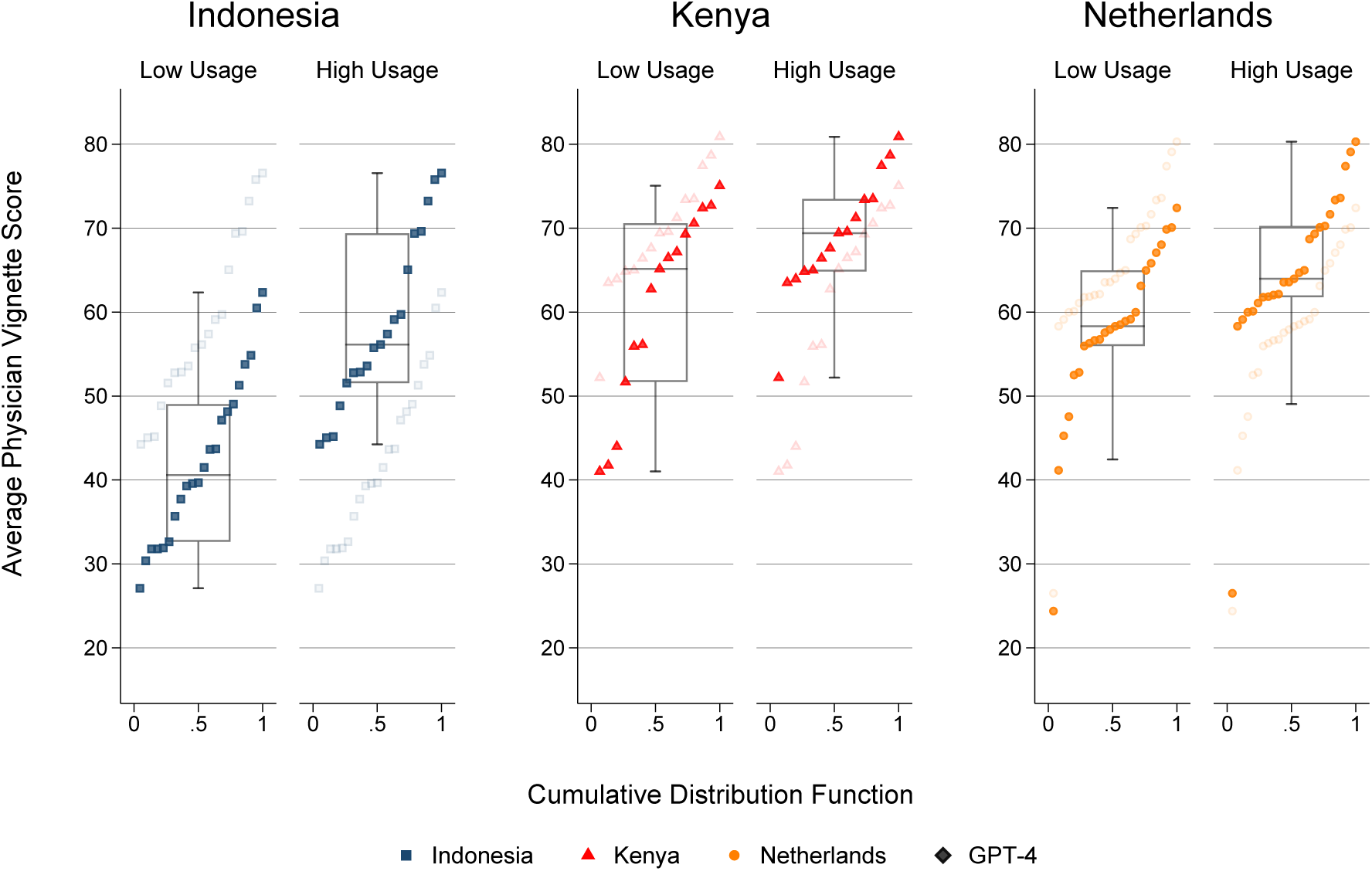
Comparison of Average Physician Vignette Scores Across Countries for High and Low within experiment LLM Usage *Note:* The figure shows average physician vignette scores for the with access group in the three countries, grouped by low and high within experiment usage of LLMs. Boxplots indicate score distributions: the boxes span the interquartile range (IQR), and whiskers extend to the minimum or maximum values within 1.5×IQR. Individual scores are shown as dots or triangles, and overlaid cumulative distribution functions (CDFs) provide a smoothed view of score distributions within each group.

## 3 Discussion

In three randomized controlled trials conducted in Indonesia, Kenya, and the Netherlands, we found that providing physicians with access to GPT-4o improved their clinical vignette scores by 10.7% in Indonesia, 18% in Kenya, and 7.2% in the Netherlands compared to physicians without access. Our primary results are in line with similar research investigating the effects of LLM access on physician performance, which demonstrate potential for augmentation [18, 25, 32]. Below, we place these results in context by highlighting four key contributions: (1) cross-country generalizability, (2) distributional impacts of LLM access, (3) compliance to the intervention, and (4) a potential mechanism via enhanced guideline adherence.

First, we demonstrate the effectiveness of LLM access in different settings. Medical education and clinical reasoning contexts can vary from country to country [20, 31] and thus we cannot expect, a priori, that access could directly improve physician performance in different settings. By conducting our trial in three economically and geographically diverse settings, each using the same four globally prevalent primary-care vignettes, we provide evidence that access to an LLM can improve clinical performance in all three contexts. These findings support the nascent literature [18, 25, 32] and demonstrate that the findings are generalizable to different countries and contexts.

Second, we examined the distributional effects of LLM access within and across our sample selections. In Kenya, physicians with access to an LLM were more homogeneous in their vignette performance than physicians without access, suggesting that LLM access could reduce physician performance variation. In contrast, physicians with access in Indonesia were more dispersed, suggesting that the effect of LLM access could lead to greater dispersion in certain settings. We hypothesize that this divergence stems from differential LLM uptake: LLM usage was higher and more consistent in Kenya than in Indonesia or the Netherlands, and usage correlated positively with performance. This suggests that the impact of LLMs may depend not only on access but also on how actively and effectively they are used. These findings demonstrate the potential of LLMs to reduce inequalities in clinical performance. However, they may inadvertently exacerbate disparities due to differential usage across physicians.

Moving to cross-country comparisons we observed significant differences in mean scores for all pairwise comparisons for physicians without access to an LLM, with the Dutch sample achieving the highest mean performance. For physicians with access to an LLM, the performance gap between Kenya and the Netherlands reversed, with Kenyan physicians scoring marginally higher than Dutch physicians. Analysis in Supplement 2 suggests that the larger treatment effect in Kenya was in part driven by an improved performance in the non-internal medicine subgroup. This is in line with other studies demonstrating that suggest LLMs boost performance among lower skilled or tenured individuals [12, 41, 49]. Further research should investigate the links between skill levels, experience, expertise, and large language models. Comparing Indonesia with the Netherlands, we also observed a reduction in the gap. These results suggest that LLMs could be an equalizer in performance disparities across countries, as well as within a country. An exploratory observation from our data suggests that a small subset of physicians in the LLM access groups in each country outperformed both the without access group and the LLM. While this finding must be interpreted cautiously given the lack of repeat LLM observations, it raises the possibility that certain users were able to extract disproportionate value from LLM access. This may reflect individual differences in how physicians engage with the technology. For example, some users may function as effective Bayesian updaters, incorporating LLM-generated recommendations into their reasoning while preserving critical judgment [4]. Others may benefit from more skillful prompting, eliciting higher-quality outputs that enhance decision-making even when incorporated passively. Complementing this, we found that the worst performers in the Netherlands are in the with access group. This can partially be attributed to low usage, however there may also be behavioral aspects. Humans suffer from automation biases whereby system advice can be uncritically taken [2, 57]. This can lead to erroneous information being incorporated into the human decision making process, and thus human-AI collaboration may degrade overall performance relative to humans or AI alone [4, 30]. Understanding the procedural, behavioral and cognitive factors that enable effective use is an important avenue for future research [18]. Third, we examined differential usage patterns of the LLM, highlighting that those who used the LLM in more steps in general performed better across all countries in our sample. We also found higher overall usage in Kenya than in Indonesia or the Netherlands. These results suggest that merely providing access may not be sufficient to improve physician performance and that active use may need to be encouraged. However, as with the difference between the with and without access groups, there is overlap between high usage and low usage groups, suggesting that high usage alone is not a determinant for better performance. Understanding why some physicians may chose to use an LLM and others not, and why, is an imperative question.

Fourth, we highlight a mechanism through which LLM access may improve physician performance. Our study uses rubrics that reflect context specific, evidence-based best practice. Thus, performance on our test is a measure of adherence to clinical best practice. Our results suggest that GPT-4o responses codify and retrieve guideline-based steps that physicians then incorporate into their answers. In effect, LLM access appears to help physicians follow codified best practice from investigations and research, through diagnosis and to patient management. LLM access could help tackle well established deficiencies in physician adherence to best practice guidelines [1, 16, 38, 42, 43]. Lack of adherence has led to the 60-30-10 problem: 60% of care on average is in line with evidence- or consensus-based guidelines, 30% is some form of waste or of low value, and 10% is harm [10]. Further research should investigate how LLMs codify and interact with guideline driven care, especially when guidelines across countries may differ.

### Our study contains a number of limitations

First, we cannot assume our results will hold for all medical conditions, either in primary care or otherwise. Our case selection focused on globally prevalent conditions with well-established evidence-based best practice guidelines. Differences could arise either from changing physician or LLM behavior. Physicians tend to use the non-analytical or heuristic approach in their clinical reasoning when dealing with common cases, and will switch to a more deliberate analytical approach on more complicated cases [55]. This could then change how they interact with LLMs. For LLM behavior, we are unable to test for bias in the LLM training data. Future work should test LLM performance on more regionally specific case selections, especially focusing on discovering biases between the Global North and South. Similarly, racial and ethnic biases in the LLM may arise that could affect cross-country variations in quality of care [23]. Further, as our cases are globally prevalent and well covered in guidelines, it can be assumed that LLMs will have codified the relevant knowledge. In related research, we demonstrate heterogeneous effects of LLM access for a more challenging and rare patient case with positive effects for diagnostic reasoning, no differential effect between challenging and standard cases for investigative reasoning, and no effect of access on management reasoning [35].

Second, cross-country differences could be attributed to changes in the rubric across Kenya, Indonesia, and the Netherlands and English proficiency across countries. To address this in Supplement 4, Table 4.2, we demonstrate that cross-country gaps are reduced but not eliminated when only using rubric items found in all 3 context specific rubrics. Regarding language differences, all physicians within Indonesia are expected to have achieved a certain level of English proficiency test, such as TOEFL during the entrance exam.

Third, participants in the without access group were not provided with traditional resources, such as clinical guidelines or web search. Access to such resources was restricted for two primary reasons: to ensure a baseline comparison that was not driven by differing resources in each context and to avoid contamination as LLMs were widely available on the internet and incorporated into search such as Google. Evidence from similar studies is mixed, with one showing that LLM access groups out-perform those with traditional resources [25], while another shows no difference [24]. In both cases however, LLM access does not reduce average performance.

Finally, this study was conducted in computer laboratories, a more ideal setting that is minimally influenced by external factors. In a real clinical setting, decision making could be influenced by the unavailability of diagnostic tools and treatments [8], insurance rules [7], and other types of interruption [51], decreasing practitioner efficiency.

## 4 Methods

### 4.1 Ethical Approval

The study was reviewed and approved by review boards of the participating universities (University of Indonesia, Aga Khan University Nairobi, and Maastricht University). Written informed consent was obtained from participants preceding enrollment and randomization. Participants were not compensated for participating in this study. We follow the CONSORT reporting guideline for randomized trials. The study protocol is available in Supplement 1. The study design was preregistered April 17, 2024 at AEA RCT Registry (RCT ID: AEARCTR-0013399).

### 4.2 Study Design

Participants completed 4 clinical vignettes, designed to simulate patient-physician consultations. We follow the structure of clinical performance and value vignettes that have previously been used to measure physician clinical performance in diverse global settings [13, 45–48]. Our vignette design follows 9 stages:

1. Presenting Problem & Initial Differential Diagnosis
2. Asking about patient history
3. Additional Differential Diagnosis
4. Listing Physical Exams
5. Differential Diagnosis
6. Additional diagnostic (Lab) Tests
7. Final Differential Diagnosis
8. Medication
9. Follow-Up/Advice

Information was provided sequentially and at each stage participants listed the actions they would take. All clinical vignettes were presented to participants in English, participants were allowed to respond either in English or their own language. All medical education is delivered in English in Kenya. In Indonesia, participants i.e residents are required to take the TOEFL test during their enrollment as resident. In the Netherlands, physicians are expected to be able to read English to at least a B2 level, in order to keep on top of developments in professional literature.

Participants were randomly assigned either to the control or intervention group using simple randomization and asked to complete the vignettes in an online environment. Intervention group participants were given access to an LLM (GPT-4o) via the OpenAI API through an interface developed by Maastricht University. This interface was integrated into the online Qualtrics environment. There was no affiliation with OpenAI. The intervention group were instructed that they could optionally use the LLM, and were provided with prompting instructions. Control group participants were not provided with any specific technology or additional resources and were asked not to use internet search.

### 4.3 Participants

Our target group consists of residents in family and internal medicine. Recruitment was supplemented with attending physicians in internal medicine in the Netherlands, and first-year residents in other specialties (i.e., surgery, anesthesiology, and pediatrics) or post-internship pre-residency medical officers, referred to as Senior House Officers, in Kenya. Participants were recruited through the university networks of the three participating universities (Maastricht University, Universitas Indonesia, Aga Khan University). Written informed consent was obtained from participants preceding enrollment and randomization. Participants were not compensated for participating in this study. Sessions were organized in controlled environments: computer labs in Indonesia and the Netherlands, and educational consultation rooms in Kenya.

### 4.4 Vignette Case Development

The selected conditions (cardiovascular, respiratory, musculoskeletal disease, fatigue diseases, and infectious disease) are globally prevalent and can be diagnosed and treated in primary care without the need for expensive treatments, advanced technology, or specialized care. They are supported by established and developed evidence-based best practice guidelines. Each case was developed for this trial by a team of experienced clinical vignette developers, with representatives from each country. To assess the validity of the case selection, participants were asked whether the patient cases presented were representative of those typically encountered in clinical practice. Participants across all three countries reported high levels of agreement: 89% in Indonesia, 92% in Kenya, and 94% in the Netherlands. No significant differences were observed between the control and intervention groups.

### 4.5 Rubric Development

Rubrics were developed based on evidence-based best practices, drawing on comprehensive national guidelines i.e. Dutch College of General Practitioners (NHG) and the UK’s National Institute for Health and Care Excellence (NICE). These best-practice rubrics were then adapted by domain experts in each country to reflect local clinical contexts. Preplanned sensitivity analyses address cross-country differences in the rubric, reported in Supplement 4, Table 4.1. Each rubric item was applied a weighting based on its clinical significance, using a standardized scale of 0.33, 0.5, or 1.

### 4.6 Response Grading & Primary Outcome generation

Open-ended participant responses were independently graded by two reviewers using locally adapted rubrics. Graders were recruited through university networks and included recently graduated internal medicine physicians in Indonesia (n=11) and Kenya (n=4), and final-year medical students in the Netherlands (n=8).

Each rubric item was assessed as present (1) or absent (0) by the reviewers. The primary outcome was calculated as the weighted sum of present items divided by the weighted sum of total possible items, creating a percent correct score per vignette. This was then averaged across participants. Pre-planned sensitivity analyses were conducted with scores generated at the vignette level using linear regressions and mixed effects models (see Supplement 5, Table 4.1 & 4.2).

We calculated Cohen’s kappa for all rubric items, which yielded a value of 0.71 (Indonesia: 0.68; Kenya: 0.67; Netherlands: 0.77 ), indicating substantial agreement. We also calculated a one-way random-effects intraclass correlation coefficient (ICC) for our primary outcome at the vignette level. The calculated ICC was 0.96 (95% CI: .957 to .966; p<.001) indicating excellent agreement in composite scores between graders.

To address disagreements in assessment, we recruited a third expert reviewer in each country to adjudicate on all disagreements. Sensitivity analyses were conducted including adjudicated results and are reported in Supplement 3, Table 3.1. Further, in Supplement 5, Table 5.3, we report results for models run removing vignettes with the highest distance between reviewer assessment (*>*10% difference between the reviewers in their score assessment).

### 4.7 Statistical Analysis

A power analysis was conducted to determine the appropriate sample size for detecting meaningful effects in the randomized controlled trial. Using vignette parameters derived from the literature [47], including a mean vignette score of 71 and standard deviation of 5.4, a two-means clustered power analysis was implemented using Stata 17. With an assumed intra-cluster correlation of 0.9 and targeting a power of 0.8, the analysis suggested that a minimum of 50 participants would be sufficient to detect a lower-bound effect size of 4.8%, based on literature estimates [9, 28–30].

Analyses were conducted using Stata/SE 17.0. All analyses were conducted using linear regressions, with cluster robust standard errors pooled by participant.

## Data Availability

All data produced in the present study are available upon reasonable request to the authors

## Notes

### Competing Interest Statement

The authors have declared no competing interest.

### Clinical Trial

AEARCTR-0013399

### Funding Statement

This study was funded INRIA on behalf of the Global Partnership on Artificial Intelligence

### Author Declarations

Ethics committee of Maastrict University gave ethical approval for this work Ethics committee of Aga Khan University Nairobi gave ethical approval for this work Ethics committee of Universitas Indonesia gave ethical approval for this work

### Summary of Updates

Fixed typo in author list on printed article from Anastacia Mbithi to Annastacia Mbithi

