## Supplementary material for "Impact of LLM Assistance on Physician Decision-Making: A Multi-Country Randomized Controlled Trial^∗^": Online Appendix

### Supplementary Materials

### Contents


**1 Study Protocol** **2**

**2 Kenya Sub-Group Analysis** **5**

**3 Sensitivity Analyses 1: Third Review** **9**

**4 Sensitivity Analyses 2: Rubric & Step Adjustment** **11**

**5 Sensitivity Analyses 3: Vignette Level** **14**

### Supplement 1: Study Protocol

#### Title

**How Large Language Models Can Affect Clinical Reasoning: A Randomized Controlled Trial on Physicians' Use of Large Language Models**

- **Design:** Parallel-group randomized controlled trial using a superiority framework.
- **Setting:** Clinical vignettes administered in computer labs at universities in Kenya, Indonesia, and the Netherlands.
- **Procedure:** Physicians completed clinical vignettes—hypothetical but viable patient scenarios—via an online Qualtrics survey. Half of the participants were randomly assigned an AI assistant (ChatGPT-4 via OpenAI API integrated into Qualtrics). The other half received no AI assistance.

**Clinical Vignette Structure:**

1. Initial differential diagnosis (limited information)
2. Patient history
3. Additional differential diagnoses
4. Physical examination
5. Third differential diagnosis
6. Further investigations
7. Final differential diagnosis
8. Treatment or medication
9. Follow-up advice

#### Trial Registration

- **Registry:** AEA Registry
- **Trial ID:** AEARCTR-0013399
- **Title:** *The Big Unknown: A Journey into Generative AI's Transformative Effect on Professions, starting with Medical Practitioners*
- **Registration Link:** `https://www.socialscienceregistry.org/trials/13399`
- **Initial Registration:** April 17, 2024
- **First Published:** April 25, 2024, 11:44 AM EDT

#### Study Design

- **Type:** Interventional (Clinical Trial)
- **Model:** Parallel group design, 1:1 allocation, superiority framework
- **Randomization:** Simple randomization using Qualtrics' randomizer (Evenly Present Elements)
- **Blinding:** Participants not blinded; graders were blinded to condition and identity
- **Setting:** University computer labs in Kenya, Indonesia, and the Netherlands
- **Eligibility:** Resident physicians (internal/family medicine), first-year residents in other specialties, and post-internship, pre-residency physicians with knowledge of primary care
- **Sample Size:** Indonesia: 81; Kenya: 60; Netherlands: 108
- **Intervention:** Access to GPT-4 via custom Qualtrics interface
- **Control:** No AI assistant

#### Study Arms

**Active Comparator:** GPT-4 via custom-built interface embedded in Qualtrics (iFrame)

**No Intervention:** No additional resources

#### Outcome Measures

**Grading:** In each country, two independent graders assessed responses against a rubric. Graders were randomly assigned from a pool (Kenya: 4, Indonesia: 8, Netherlands: 11). Disagreements were resolved by a third, more experienced reviewer. All final scores required agreement between at least two reviewers. Resolved disagreements are used for sensitivity analysis

**Primary Outcome:** Percent correct score (rubric-based). Calculated as the weighted number of rubric items marked as present, divided by the total possible.

**Note on Step 3:** Many participants left Step 3 blank as the question was asked as "List any additional diagnoses". To adjust, rubric items marked in Step 1 were counted toward Step 3 if relevant. Sensitivity analyses will test robustness by removing Step 3 entirely.

#### Power Analysis

Using `Stata/SE 17.0`, a two-means clustered power analysis was performed:

- **Baseline:** Mean vignette score = 71, SD = 5.4 (Peabody et al., 2000)
- **ICC:** 0.9 (participants as clusters, vignettes as observations)
- **Target Power:** 80%
- **Minimum Sample:** 50 participants required to detect a 4.8% effect size (based on literature: Bien et al., 2018; Han et al., 2020; Jain et al., 2021; Jussupow et al., 2021)

#### Statistical Analysis

- Descriptive statistics (means, SD, medians, IQRs) for continuous variables; proportions for categorical variables
- OLS regressions per country at participant level, with clustering at the participant level
- Separate analysis for cognitively difficult vignette

**Preplanned Sensitivity Analyses:**

- Mixed-effects model with participant, vignette, and question intercepts
- Only rubric items shared across all countries
- Cases with third-reviewer adjudication only
- Exclude vignettes with >10% grader disagreement
- Remove Step 3 from all outcomes

### Supplement 2: Kenya Sub-Group Analysis

In this supplement, we assess the impact on our main findings for Kenya of the unbalanced sample distribution between two subgroups in our data: the Internal Medicine residents and residents specialising in other areas, or pre-residency physicians (referred to as non-specialists). We do so because the unbalanced sample distribution across intervention and control is potentially confounding for a number of conclusions we draw in the paper. This is driven by the fact that internal medicine residents may have been exposed to training on guidelines that cover the four patient conditions used in this study.

We explore the impact in Supplement Figure 2.1. In this study, we argue that LLM access contributed to increased homogeneity in the sample of Kenyan physicians with access to an LLM. The evidence we present in this supplement suggests that within the intervention group Internal Medicine specialists and non-specialists exhibit similar performance outcomes, with comparable medians and interquartile ranges (IQRs). By contrast, the group without LLM access show substantial differences between the two subgroups of Internal Medicine specialists and non-specialists, This indicates that LLM access may reduce performance differences/variability between Internal Medicine specialists and non-specialists.

Supplement Table 2.1 reports the estimates of Table 1 in the main manuscript. Panel A reports on effects across three groups: the full sample, the Internal Medicine Specialists, and the non-specialist subgroup. As is to be expected, the intervention effect is larger for those in the non-specialist sub group, however, we still see a significant effect for the Internal Medicine subgroup. The estimated mean difference for the Internal Medicine subgroup in Kenya is 10.6% (95% CI:3.4-17.9, p=0.006) which is similar in magnitude to the effect found in Indonesia of 10.7%(95% CI: 5.7 to 15.7, p<0.001) and the Netherlands 7.2% (95% CI: 3.7 to 10.7, p<0.001).

Panel B reports the difference across the two subgroups. This shows that the gap between the two subgroups for physicians with access is 9.8% (95% CI: 0-19.7, p=0.004) smaller than for those without. Further, Kolomgorov-Smirnov tests confirm that the distributions of physicians specialising in Internal Medicine and the non-specialist subgroup are statistically significantly different for physicians without access to an LLM, while no such difference can be detected across the subgroup for the group with access . Taken together, these results are consistent with the hypothesis that LLM access reduces performance variability among physicians in Kenya.

We explore this further in Figure 2.2, which disaggregates the Kenyan intervention group into Internal Medicine and non-specialised subgroups and further stratifies each by high and low LLM usage. The figure shows that most of the outliers belong to the low-usage group across both subgroups. This pattern further supports our hypothesis that LLM access decreases performance variability in the Kenya sample. The increased homogeneity in the intervention group is driven by active usage of the LLM.

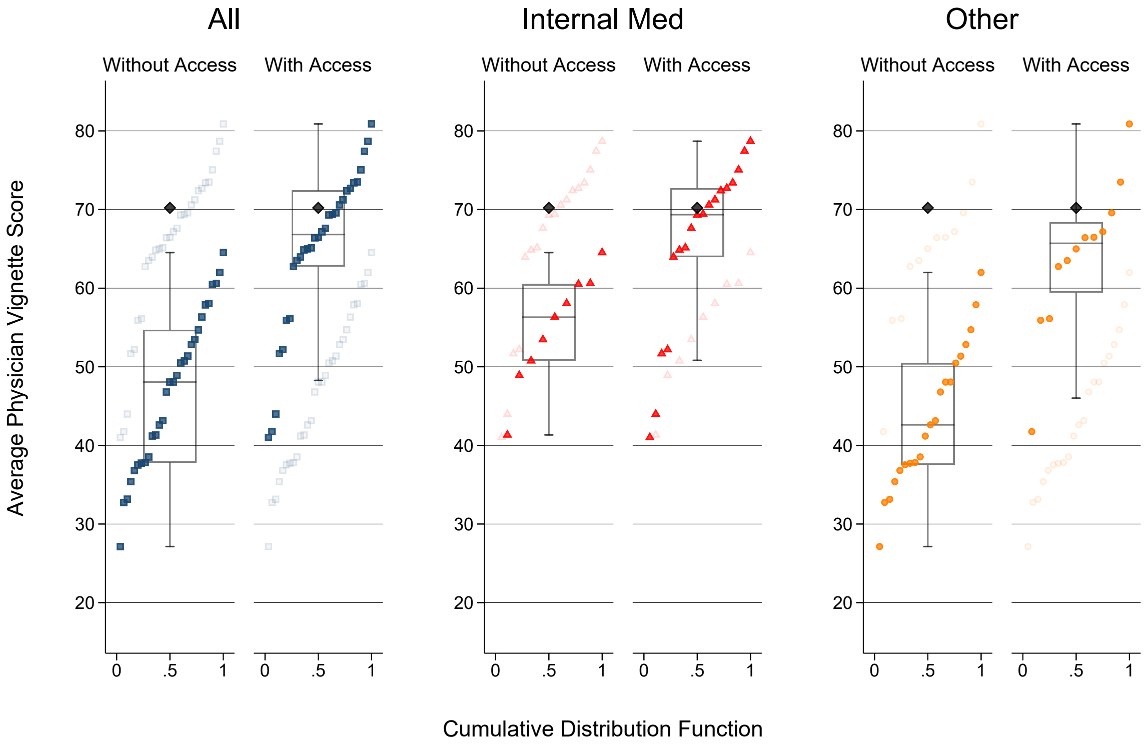


Figure 2.1: Comparison of Average Physician Vignette Scores Across Sub-Samples With and Without LLM Access

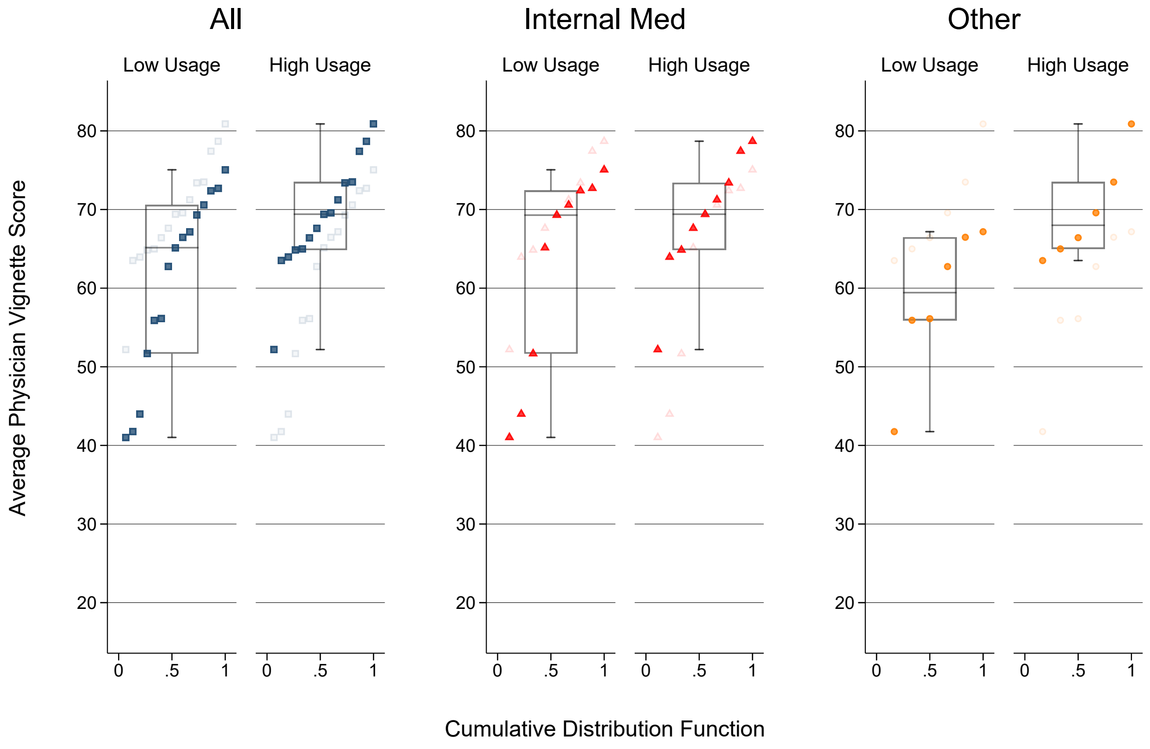


Figure 2.2: Comparison of Average Physician Vignette Scores Across Sub-Samples for High Usage and Low Usage

Table 2.1: Comparison of Average Physician Vignette Scores Across Specialisation With and Without LLM access

| | | Without Access | With Access | Mean Differences (CI 95%) |
|---|---|---|---|---|
| **Panel A-Within Sample Effects** | | | | |
| All | | | | |
| | Observations | 30 | 30 | |
| | Mean(SD) | 47(10) | 65(10.4) | 18(12.7-23.2), p<0.001 |
| | Median(IQR) | 48.1(37.8-54.7) | 66.8(62.7-72.4) | |
| | Min-Max Difference (Min-Max) | 37.4(27.1-64.5) | 39.9(41-80.9) | |
| | Kolmogorov-Smirnov | | | p<0.001 |
| Internal Med | | | | |
| | Observations | 9 | 18 | |
| | Mean(SD) | 54.9(7.1) | 65.6(11.1) | 10.6(3.4-17.9), p=0.006 |
| | Median(IQR) | 56.3(50.8-60.5) | 69.3(64-72.7) | |
| | Min-Max Difference (Min-Max) | 23.2(41.3-64.5) | 37.7(41-78.7) | |
| | Kolmogorov-Smirnov | | | p=0.002 |
| Other | | | | |
| | Observations | 21 | 12 | |
| | Mean(SD) | 43.6(9.1) | 64.1(9.8) | 20.5(13.5-27.5), p<0.001 |
| | Median(IQR) | 42.6(37.5-50.5) | 65.7(59.4-68.4) | |
| | Min-Max Difference (Min-Max) | 34.9(27.1-62) | 39.1(41.8-80.9) | |
| | Kolmogorov-Smirnov | | | p<0.001 |
| **Panel B - Cross-Sample Effects** | | | | |
| Internal Med - Other | | | | |
| | Mean (95% CI) | -11.3(-17.6–5),p<0.001 | -1.5(-9.4-6.4),p=0.70 | 9.8(0-19.7),p=0.004 |
| | Kolmogorov-Smirnov | p=0.01 | p=0.26 | |

### Supplement 3: Sensitivity Analyses 1: Third Review

Table 3.1: Primary outcome is with decisions of third expert reviewer

| | | Without Access | With Access | Mean Differences (CI 95%) |
|---|---|---|---|---|
| **Panel A-Within Country Effects** | | | | |
| Indonesia | | | | |
| | Observations | 40 | 41 | |
| | Mean(SD) | 40.9(9.9) | 52.7(14) | 11.8(6.5-17.2), p<0.001 |
| | Median(IQR) | 40.7(35.8-47.6) | 52.6(42.6-60.4) | |
| | Min-Max Difference (Min-Max) | 42.2(18.6-60.8) | 53.8(27.8-81.6) | |
| | Kolmogorov-Smirnov | | | p<0.001 |
| Kenya | | | | |
| | Observations | 30 | 30 | |
| | Mean(SD) | 46.7(10) | 65.5(11) | 18.8(13.3-24.2), p<0.001 |
| | Median(IQR) | 47.9(38.1-54.7) | 67.3(62.3-71.8) | |
| | Min-Max Difference (Min-Max) | 39.3(25.9-65.2) | 42.7(39.6-82.3) | |
| | Kolmogorov-Smirnov | | | p<0.001 |
| Netherlands | | | | |
| | Observations | 58 | 50 | |
| | Mean(SD) | 54.8(6.7) | 63.1(11.5) | 8.3(4.6-12), p<0.001 |
| | Median(IQR) | 56.4(51.1-59.8) | 64(59.1-70.9) | |
| | Min-Max Difference (Min-Max) | 28.9(38.1-67) | 58(22.8-80.8) | |
| | Kolmogorov-Smirnov | | | p<0.001 |
| **Panel B - Cross-National Effects** | | | | |
| Indonesia - Kenya | | | | |
| | Mean (95% CI) | 5.8(1-10.6),p=0.02 | 12.8(6.8-18.7),p<0.001 | 6.9(-0.6-14.5),p=0.07 |
| | Kolmogorov-Smirnov | p=0.08 | p<0.001 | |
| Indonesia - Netherlands | | | | |
| | Mean (95% CI) | 13.9(10.3-17.4),p<0.001 | 10.4(5-15.8),p<0.001 | -3.5(-9.9-2.9),p=0.28 |
| | Kolmogorov-Smirnov | p<0.001 | p<0.001 | |
| Kenya - Netherlands | | | | |
| | Mean (95% CI) | 8.1(4.1-12.1),p<0.001 | -2.4(-7.5-2.8),p=0.36 | -10.4(-16.9- -4),p=0.002 |
| | Kolmogorov-Smirnov | p=0.002 | p=0.40 | |

### Supplement 4: Sensitivity Analyses 2: Rubric & Step Adjustment

Table 4.1: Results with consistent rubric across countries

| | | Without Access | With Access | Mean Differences (CI 95%) |
|---|---|---|---|---|
| **Panel A-Within Country Effects** | | | | |
| Indonesia | | | | |
| | Observations | 40 | 41 | |
| | Mean(SD) | 41.7(10.2) | 52.6(13.5) | 10.9(5.6-16.2), p<0.001 |
| | Median(IQR) | 41.2(36.6-47.2) | 52.8(42.9-60.7) | |
| | Min-Max Difference (Min-Max) | 43.9(18.6-62.5) | 50.8(29.1-79.9) | |
| | Kolmogorov-Smirnov | | | p<0.001 |
| Kenya | | | | |
| | Observations | 30 | 30 | |
| | Mean(SD) | 49.2(10.2) | 67.6(10.7) | 18.4(13-23.8), p<0.001 |
| | Median(IQR) | 50.1(39.7-57.9) | 69.9(64.7-74.9) | |
| | Min-Max Difference (Min-Max) | 36.5(30.1-66.6) | 40.5(43.3-83.8) | |
| | Kolmogorov-Smirnov | | | p<0.001 |
| Netherlands | | | | |
| | Observations | 58 | 50 | |
| | Mean(SD) | 54.4(6.5) | 61.6(10.9) | 7.2(3.7-10.7), p<0.001 |
| | Median(IQR) | 55.7(50.9-59.7) | 62.2(58.2-69) | |
| | Min-Max Difference (Min-Max) | 25.5(39.1-64.6) | 56(24.5-80.5) | |
| | Kolmogorov-Smirnov | | | p<0.001 |
| **Panel B - Cross-National Effects** | | | | |
| Indonesia - Kenya | | | | |
| | Mean (95% CI) | 7.5(2.6-12.4),p=0.003 | 15(9.2-20.7),p<0.001 | 7.5(0-15),p=0.05 |
| | Kolmogorov-Smirnov | p=0.02 | p<0.001 | |
| Indonesia - Netherlands | | | | |
| | Mean (95% CI) | 12.7(9.1-16.3),p<0.001 | 9(3.8-14.2),p<0.001 | -3.7(-10-2.6),p=0.25 |
| | Kolmogorov-Smirnov | p<0.001 | p<0.001 | |
| Kenya - Netherlands | | | | |
| | Mean (95% CI) | 5.2(1.2-9.3),p=0.01 | -6(-10.9- -1),p=0.02 | -11.2(-17.6–4.9),p<0.001 |
| | Kolmogorov-Smirnov | p=0.01 | p=0.005 | |

Table 4.2: Results with Vignette Step 3 removed

| | | Without Access | With Access | Mean Differences (CI 95%) |
|---|---|---|---|---|
| **Panel A-Within Country Effects** | | | | |
| Indonesia | | | | |
| | Observations | 40 | 41 | |
| | Mean(SD) | 38.4(10) | 49.2(13.4) | 10.7(5.5-16), p<0.001 |
| | Median(IQR) | 38.4(33.4-43.5) | 48.2(38.3-56.6) | |
| | Min-Max Difference (Min-Max) | 42(15.7-57.7) | 51.8(24.8-76.6) | |
| | Kolmogorov-Smirnov | | | p<0.001 |
| Kenya | | | | |
| | Observations | 30 | 30 | |
| | Mean(SD) | 46.4(10.1) | 64.6(10.8) | 18.2(12.8-23.6), p<0.001 |
| | Median(IQR) | 46.9(37.4-54.4) | 65.6(62.4-72.1) | |
| | Min-Max Difference (Min-Max) | 37.8(26.6-64.4) | 39.6(40.5-80.1) | |
| | Kolmogorov-Smirnov | | | p<0.001 |
| Netherlands | | | | |
| | Observations | 58 | 50 | |
| | Mean(SD) | 52.4(6.6) | 60(11.1) | 7.6(4.1-11.2), p<0.001 |
| | Median(IQR) | 53.3(48.8-57.9) | 60.4(56.3-67) | |
| | Min-Max Difference (Min-Max) | 25.1(38-63.1) | 55.8(22.4-78.2) | |
| | Kolmogorov-Smirnov | | | p<0.001 |
| **Panel B - Cross-National Effects** | | | | |
| Indonesia - Kenya | | | | |
| | Mean (95% CI) | 8(3.1-12.8),p=0.002 | 15.4(9.7-21.1),p<0.001 | 7.4(0-14.9),p=0.05 |
| | Kolmogorov-Smirnov | p=0.008 | p<0.001 | |
| Indonesia - Netherlands | | | | |
| | Mean (95% CI) | 14(10.4-17.6),p<0.001 | 10.9(5.7-16.1),p<0.001 | -3.1(-9.4-3.2),p=0.33 |
| | Kolmogorov-Smirnov | p<0.001 | p<0.001 | |
| Kenya - Netherlands | | | | |
| | Mean (95% CI) | 6(2-10),p=0.004 | -4.5(-9.5-0.5),p=0.08 | -10.5(-16.9- -4.2),p=0.001 |
| | Kolmogorov-Smirnov | p=0.01 | p=0.01 | |

### Supplement 5: Sensitivity Analyses 3: Vignette Level

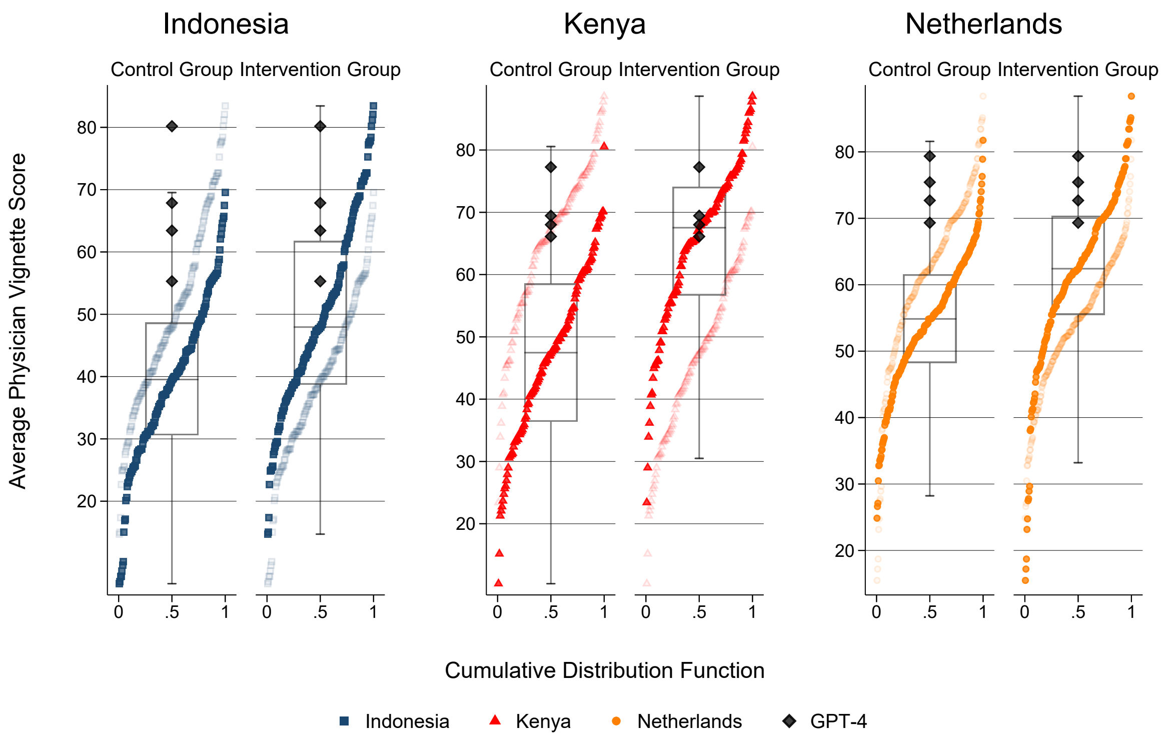


Figure 5.1: Boxplots at Vignette Level

Table 5.1: Results at the Vignette Level

| | | Without Access | With Access | Mean Differences (CI 95%) |
|---|---|---|---|---|
| **Panel A-Within Country Effects** | | | | |
| Indonesia | | | | |
| | Observations | 160 | 164 | |
| | Mean(SD) | 39.2(13.3) | 49.9(15.3) | 10.7(5.7-15.7), p<0.001 |
| | Median(IQR) | 39.5(30.6-48.7) | 47.9(38.7-61.8) | |
| | Min-Max Difference (Min-Max) | 60.4(7.1-67.5) | 67.1(15-82.1) | |
| | Kolmogorov-Smirnov | | | p<0.001 |
| Kenya | | | | |
| | Observations | 120 | 120 | |
| | Mean(SD) | 47(13.6) | 65(13.1) | 18(12.7-23.2), p<0.001 |
| | Median(IQR) | 47.5(36.4-58.6) | 67.5(56.7-74.1) | |
| | Min-Max Difference (Min-Max) | 54.9(15.2-70.1) | 58.7(29-87.7) | |
| | Kolmogorov-Smirnov | | | p<0.001 |
| Netherlands | | | | |
| | Observations | 232 | 200 | |
| | Mean(SD) | 54.2(10.1) | 61.4(13.2) | 7.2(3.7-10.7), p<0.001 |
| | Median(IQR) | 54.9(48.2-61.6) | 62.4(55.5-70.3) | |
| | Min-Max Difference (Min-Max) | 49.2(27.1-76.3) | 67(18-85) | |
| | Kolmogorov-Smirnov | | | p<0.001 |
| **Panel B - Cross-National Effects** | | | | |
| Indonesia - Kenya | | | | |
| | Mean (95% CI) | 7.8(3.1-12.5),p=0.001 | 15.1(9.6-20.6),p<0.001 | 7.3(0.1-14.4),p=0.05 |
| | Kolmogorov-Smirnov | p<0.001 | p<0.001 | |
| Indonesia - Netherlands | | | | |
| | Mean (95% CI) | 15(11.6-18.4),p<0.001 | 11.5(6.4-16.5),p<0.001 | -3.5(-9.5-2.5),p=0.25 |
| | Kolmogorov-Smirnov | p<0.001 | p<0.001 | |
| Kenya - Netherlands | | | | |
| | Mean (95% CI) | 7.2(3.2-11.1),p<0.001 | -3.6(-8.5-1.2),p=0.14 | -10.8(-17- -4.6),p<0.001 |
| | Kolmogorov-Smirnov | p<0.001 | p=0.005 | |

Table 5.2: Results using Mixed Effects model

| | | Without Access | With Access | Mean Differences (CI 95%) |
|---|---|---|---|---|
| **Panel A-Within Country Effects** | | | | |
| Indonesia | | | | |
| | Observations | 160 | 164 | |
| | Mean(SD) | 39.2(13.3) | 49.9(15.3) | 10.7(5.8-15.6), p<0.001 |
| | Median(IQR) | 39.5(30.6-48.7) | 47.9(38.7-61.8) | |
| | Min-Max Difference (Min-Max) | 60.4(7.1-67.5) | 67.1(15-82.1) | |
| | Kolmogorov-Smirnov | | | p<0.001 |
| Kenya | | | | |
| | Observations | 120 | 120 | |
| | Mean(SD) | 47(13.6) | 65(13.1) | 18(12.9-23.1), p<0.001 |
| | Median(IQR) | 47.5(36.4-58.6) | 67.5(56.7-74.1) | |
| | Min-Max Difference (Min-Max) | 54.9(15.2-70.1) | 58.7(29-87.7) | |
| | Kolmogorov-Smirnov | | | p<0.001 |
| Netherlands | | | | |
| | Observations | 232 | 200 | |
| | Mean(SD) | 54.2(10.1) | 61.4(13.2) | 7.2(3.9-10.5), p<0.001 |
| | Median(IQR) | 54.9(48.2-61.6) | 62.4(55.5-70.3) | |
| | Min-Max Difference (Min-Max) | 49.2(27.1-76.3) | 67(18-85) | |
| | Kolmogorov-Smirnov | | | p<0.001 |
| **Panel B - Cross-National Effects** | | | | |
| Indonesia - Kenya | | | | |
| | Mean (95% CI) | 7.8(3.3-12.4),p<0.001 | 15.1(9.6-20.6),p<0.001 | 7.3(0.1-14.5),p=0.05 |
| | Kolmogorov-Smirnov | p<0.001 | p<0.001 | |
| Indonesia - Netherlands | | | | |
| | Mean (95% CI) | 15(11.9-18.1),p<0.001 | 11.5(6.7-16.3),p<0.001 | -3.5(-9.2-2.2),p=0.23 |
| | Kolmogorov-Smirnov | p<0.001 | p<0.001 | |
| Kenya - Netherlands | | | | |
| | Mean (95% CI) | 7.2(3.8-10.6),p<0.001 | -3.6(-8.4-1.2),p=0.14 | -10.8(-16.6- -5),p<0.001 |
| | Kolmogorov-Smirnov | p<0.001 | p=0.005 | |

Table 5.3: Results at the Vignette Level with vignettes with more than 10% difference between reviewers removed

| | | Without Access | With Access | Mean Differences (CI 95%) |
|---|---|---|---|---|
| **Panel A-Within Country Effects** | | | | |
| Indonesia | | | | |
| | Observations | 135 | 135 | |
| | Mean(SD) | 39.4(13.3) | 49.6(15.4) | 10.5(5.3-15.6), p<0.001 |
| | Median(IQR) | 39.6(30.7-49) | 47.7(38.7-62.1) | |
| | Min-Max Difference (Min-Max) | 58.6(7.1-65.7) | 67.1(15-82.1) | |
| | Kolmogorov-Smirnov | | | p<0.001 |
| Kenya | | | | |
| | Observations | 105 | 106 | |
| | Mean(SD) | 46.8(13.9) | 64.9(13.5) | 17.9(12.3-23.6), p<0.001 |
| | Median(IQR) | 47(35.9-58.1) | 67.4(55.5-74.6) | |
| | Min-Max Difference (Min-Max) | 54.9(15.2-70.1) | 58.7(29-87.7) | |
| | Kolmogorov-Smirnov | | | p<0.001 |
| Netherlands | | | | |
| | Observations | 212 | 180 | |
| | Mean(SD) | 54(10) | 61.3(13.3) | 7.5(4-11.1), p<0.001 |
| | Median(IQR) | 54.8(48-61.1) | 62.4(55.5-70.3) | |
| | Min-Max Difference (Min-Max) | 49.2(27.1-76.3) | 67.9(17.2-85.1) | |
| | Kolmogorov-Smirnov | | | p<0.001 |
| **Panel B - Cross-National Effects** | | | | |
| Indonesia - Kenya | | | | |
| | Mean (95% CI) | 7.5(2.6-12.5),p=0.003 | 14.4(8.4-20.3),p<0.001 | 7.4(-0.1-15),p=0.05 |
| | Kolmogorov-Smirnov | p=0.001 | p<0.001 | |
| Indonesia - Netherlands | | | | |
| | Mean (95% CI) | 14.7(11.1-18.2),p<0.001 | 11.5(6.2-16.8),p<0.001 | -2.9(-9.2-3.3),p=0.36 |
| | Kolmogorov-Smirnov | p<0.001 | p<0.001 | |
| Kenya - Netherlands | | | | |
| | Mean (95% CI) | 7.3(3.1-11.4),p<0.001 | -3.3(-8.6-1.9),p=0.21 | -10.5(-17.1- -3.9),p=0.002 |
| | Kolmogorov-Smirnov | p<0.001 | p=0.01 | |
